# Genomics of Indian SARS-CoV-2: Implications in genetic diversity, possible origin and spread of virus

**DOI:** 10.1101/2020.04.25.20079475

**Authors:** Mainak Mondal, Ankita Lawarde, Kumaravel Somasundaram

**Author notes:** Contributed equally (Mainak Mondal, Ankita Lawarde).

## Abstract

World Health Organization (WHO) declared COVID-19 as a pandemic disease on March 11, 2020. Comparison of genome sequences from diverse locations allows us to identify the genetic diversity among viruses which would help in ascertaining viral virulence, disease pathogenicity, origin and spread of the SARS-CoV-2 between countries. The aim of this study is to ascertain the genetic diversity among Indian SARS-CoV-2 isolates. Initial examination of the phylogenetic data of SARS-CoV-2 genomes (n=3123) from different continents deposited at GISAID (Global Initiative on Sharing All Influenza Data) revealed multiple origin for Indian isolates. An in-depth analysis of 449 viral genomes derived from samples representing countries from USA, Europe, China, East Asia, South Asia, Oceania, Middle East regions and India revealed that most Indian samples are divided into two clusters (A and B) with cluster A showing more similarity to samples from Oceania and Kuwait and the cluster B grouping with countries from Europe, Middle East and South Asia. Diversity analysis of viral clades, which are characterized by specific non-synonymous mutations in viral proteins, discovered that the cluster A Indian samples belong to I clade (V378I in ORF1ab), which is an Oceania clade with samples having Iran connections and the cluster B Indian samples belong to G clade (D614G in Spike protein), which is an European clade. Thus our study identifies that the Indian SARS-CoV-2 viruses belong to I and G clades with potential origin to be countries mainly from Oceania, Europe, Middle East and South Asia regions, which strongly implying the spread of virus through most travelled countries. The study also emphasizes the importance of pathogen genomics through phylogenetic analysis to discover viral genetic diversity and understand the viral transmission dynamics with eventual grasp on viral virulence and disease pathogenesis.

## Introduction

A novel corona virus (SARS-CoV-2) causes acute respiratory disease (Coronavirus disease 2019; COVID-19), which was initially found in China but now it is spread all over the world (Guo et al., 2020). The total number of COVID-19 cases diagnosed so far exceeds 2.7 million worldwide as on 24^th^ April, 2020 with the number almost reaching 24,700 in India (https://www.arcgis.com/; https://www.mohfw.gov.in/). SARS-CoV-2 belongs to the family Coronaviridae. All viruses of this family are enveloped, non-segmented positive-sense RNA viruses. They all contain a large genome of approximately 30 kb in length. Coronaviruses are known to cause respiratory and intestinal infections. Coronavirus (CoV) are divided into four genera-alpha, beta, gamma and delta. While alpha and beta have been shown to infect mammals, gamma and delta infect birds and also mammals. Three highly pathogenic and large scale epidemic coronaviruses of the recent past would include SARS-CoV of 2012, which caused severe acute respiratory syndrome, MERS-CoV of 2012 which caused Middle East respiratory syndrome and SARS-CoV-2 of 2019 which causes COVID-19 (Khan et al., 2020; Zhou et al., 2020).

While the genome sequence of SARS-CoV-2 is 96.2% identical to a BatCoV RaTG13, it shares with 79.5% similarity to SARS-CoV. Based on this virus genome sequence analysis and evolutionary analysis, bat has been suspected to be the natural host of the virus origin. Since bats are not available at seafood markets of Wuhan, it is suspected that virus spread might have occurred through alternative intermediate hosts like turtles, and snakes (Guo et al., 2020). Since then, a total of 10,478 viral isolates have been sequenced and deposited online (https://www.gisaid.org/). Genome sequence analysis of viral genome between countries would help to understand the origin and also the severity of the disease process itself. There are only two reports of sequencing of very few numbers of Indian isolates of SARS-CoV-2 (Yadav et al., 2020; Sardar et al., 2020). The diagnosis of COVID-19 is largely overseen by Indian Council of Medical Research (ICMR), Government of India with diagnosis being carried out using commercial diagnostic kits in ICMR approved government and private facilities (https://www.icmr.nic.in/).

## Results and Discussion

To identify the origin of Indian isolates of SAR-CoV-2 virus, we examined the phylogenetic data from Database Features of Global Initiative on Sharing All Influenza Data (GISAID; https://www.gisaid.org/). Although the sequence information is available from 11,919 SARS-CoV-2 isolates from all continents, the phylogenetic data from 3123 samples, which included 4 Indian isolates, is only available at GISAID website because of limitation with respect to single view performance and legibility reasons (**Figure 1; Supplementary figure 1**). It is of our interest to note that there are two major clusters-Asian cluster represented by purple and related colours and European cluster represented by greenish yellow. The Indian samples, represented by arrows (black and white) cluster with both Asian cluster and European clusters. It is interesting to note that while the Indian samples with black arrows were isolated during January 2020, the other two samples with white arrows were isolated during March 2020 (see more details later).

**Figure 1.**
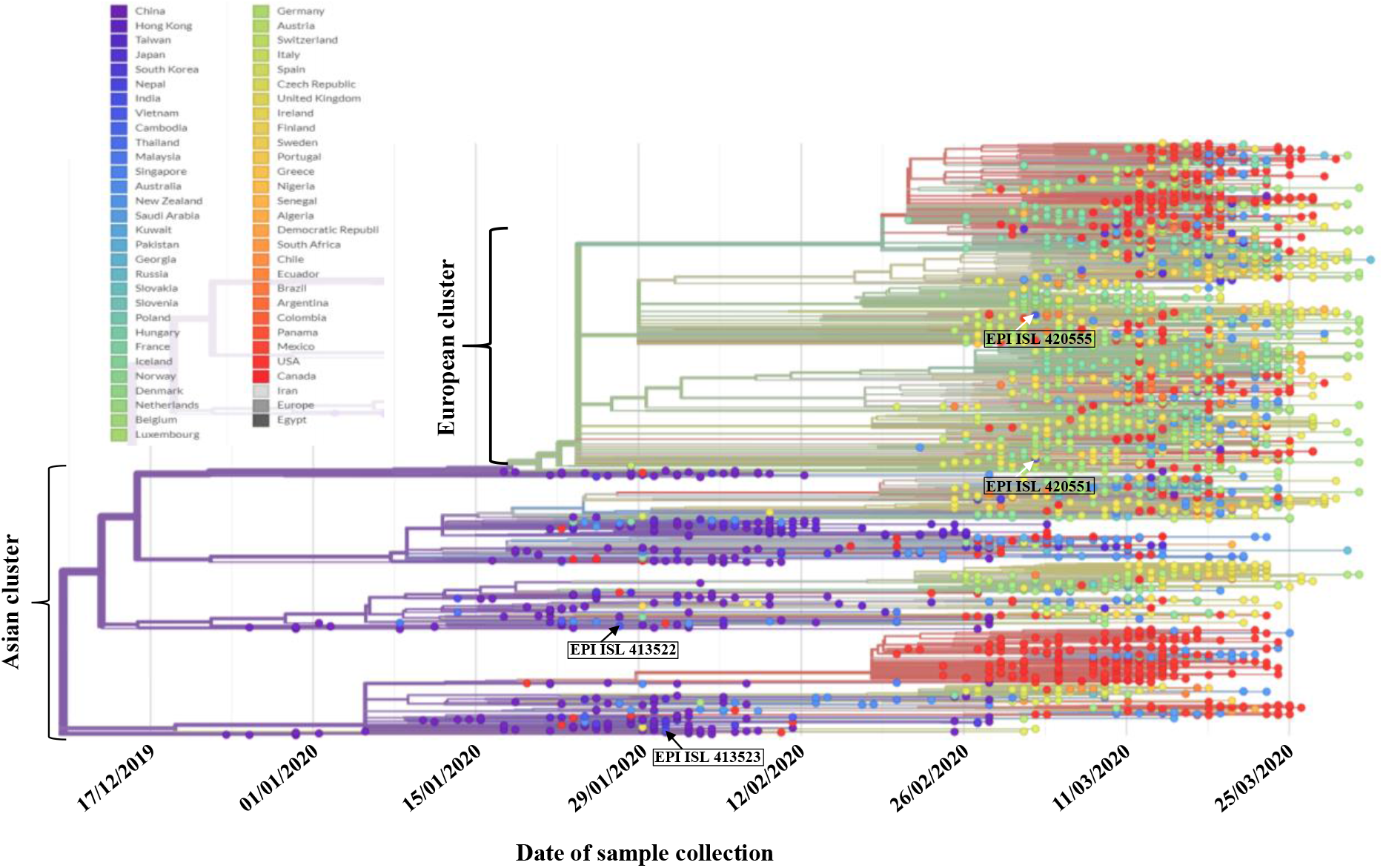
A modified view of Phylogenetic analysis (Rectangular view) of genome sequences of SARAS-CoV-2 (n=3123) taken from https://www.gisaid.org/epiflu-applications/next-hcov-19-app/. The list of countries from where samples were used is given with their colour code.

Further to precisely map the origin of Indian SARS-CoV-2 isolates, we carried out an independent phylogenetic analysis using a selected set of samples representing most regions and countries where COVID-19 infection rate is high. Our selection criteria also considered the fact that more genomes are sequenced in USA, Europe, East Asia and Oceania compared to other parts of the world. Hence, we chose a set of 449 samples derived from USA (75), Europe (80), China (75), East Asia (64), South Asia (41), Oceania (75), Middle East (11) and India (28). The analysis shows interesting features about the possible source of Indian SARS-CoV-2 samples (**Figure 2; Supplementary figure 2**). In particular, the Indian samples are divided between two clusters A and B. The cluster A consists of mostly Oceania/Kuwait samples besides a large number of Indian samples (n=13). The cluster B consists mostly of European and few numbers of Middle East/South Asian sample besides a large number of Indian samples (n=13).

**Figure 2.**
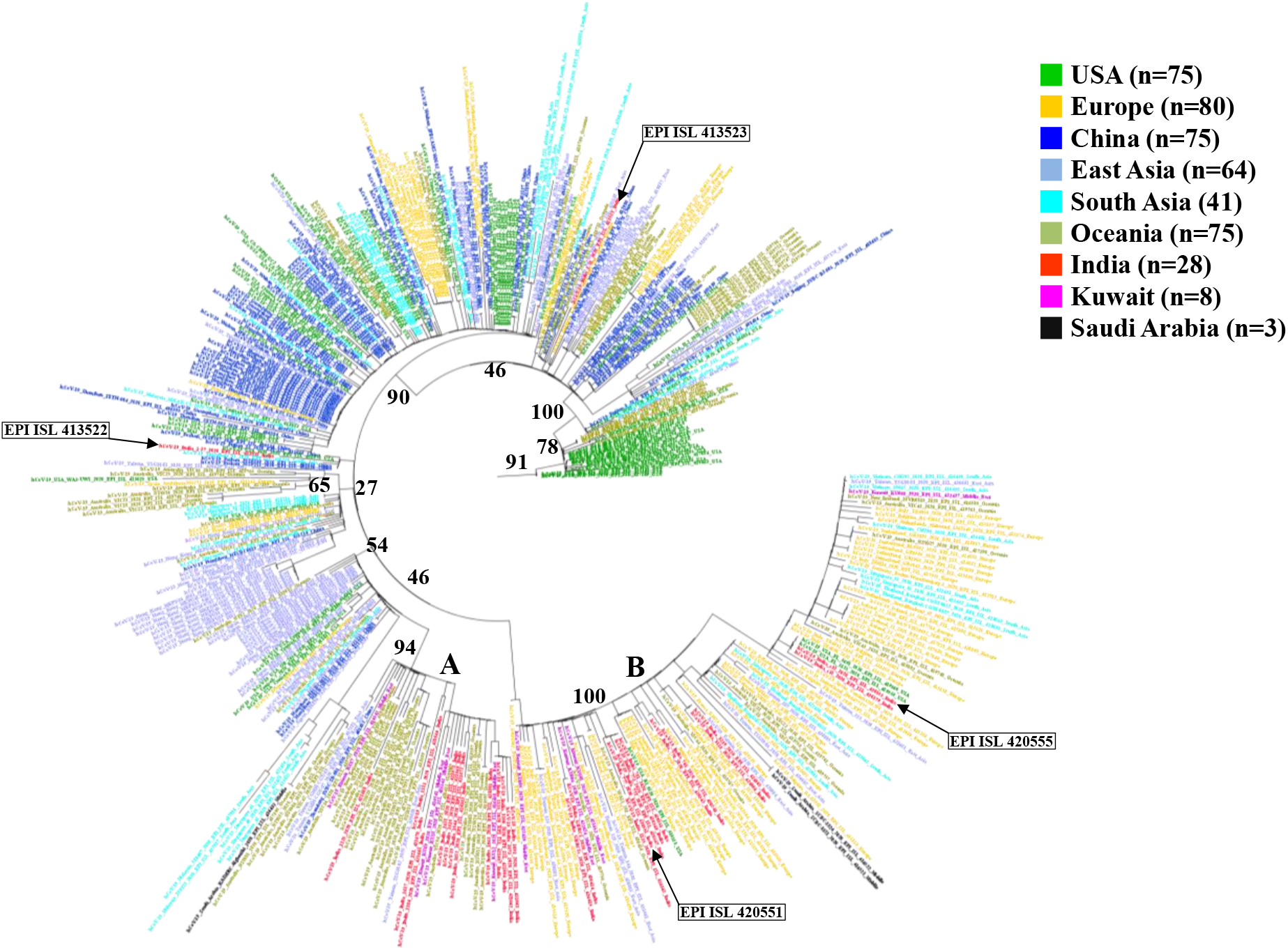
Maximum likelihood (ML) phylogenetic tree was constructed using whole genome sequences obtained from 449 individual SARS-Cov-2 viral isolates. The branch was tipped according to strains with respective country. The nodes represent bootstrap values. The taxa where coloured according to different countries. The colour code for different regions/countries is given.

The analysis revealed that most Indian SARS-CoV-2 viruses (26 out of 28) show more similarity to that of specific countries. In cluster A, Indian samples show more similarity to the viruses found in Oceania/Kuwait samples, while in the cluster B, Indian samples show more similarity to mainly European and few numbers of Middle East/South Asian samples. These results indicate that majority of Indian SARS-CoV-2 viruses have originated from Europe, Middle East, South Asia and Oceania regions. Besides these two sets of Indian SARS-CoV-2 viruses, there are two more Indian isolates (EPI_ISL_413522 and EPI_ISL_413523) which are not present in clusters A and B; these two isolates are present in other clusters which contained most samples from China and East Asia. This indicates that these two Indian SARS-CoV-2 viruses might have originated from China. Indeed, a recent study reported that these two viruses were isolated from patients who travelled from Wuhan, China (Yadav et al., 2020).

The date of sample collection probably coincides with the time of disease occurrence. The collection dates of different Indian samples provide some hint at the origin and spread of virus. The first two Indian viral isolates that were detected during January 2020 from patients who travelled from Wuhan, China showed more similarity with China/East Asia viral isolates. This conclusion is well supported by the fact that the initial outbreak of SARS-CoV-2 virus in Wuhan happened during December 2019 (Zhou et al., 2020). The remaining twenty-six Indian viral isolates, which were collected during March 2020, show more similarity with samples from Europe, Middle East, South Asia and Oceania regions. The one month delay in the occurrence of majority of Indian COVID-19 cases probably indicate the time taken for the virus to spread from China to other countries from where the Indian travellers would have contracted the virus.

It is interesting to note that two samples (EPI_ISL_420551 and EPI_ISL_420555), that are seen with European cluster in Figure 1, grouped with the cluster B, which is enriched with European and few numbers of Middle East/South Asian samples as per Figure 2. Similarly, other two samples (EPI-ISL_413522 and EPI_ISL_413523) (collected from patients who had travel history from Wuhan, China) that grouped with samples from China were also identified to be associated with the major Asian cluster in Figure 1. Thus, the results of our independent phylogenetic analysis (as per Figure 2) are matching to that of analysis done by GISAID thus increasing the confidence in our findings.

According to specific variations in different viral proteins, GISAID identified three clades of SARS-CoV-2 namely G, V and S (Elbe and Bucklan-Merrett, 2017). G clade is characterized by D614G in S protein and largely encompasses sequences from Europe. V clade is characterized by G251V in ORF3 and mostly includes Asian and European sequences. S clade is characterized by the presence of L84S in ORF8 and mostly comprises sequences from North America. Recently, a new clade of SARS-CoV-2 carrying V378I in ORF1ab has been linked to travellers returning from Iran to Australia (Eden et al., 2020). We then investigated the genomes of Indian SARS-CoV-2 viruses to find out the association between Indian samples and different clades. This analysis revealed several interesting facts (**Figure 3**). We found Indian samples does not belong to either clade V or S except the sample EPI-ISL_413522, which is one of samples derived from patient with recent travel history from Wuhan. The other Indian sample (EPI_ISL_413523) who also had travel history from Wuhan does not seem to belong to any of these clades suggesting that this virus may be more similar to original Wuhan-China virus. However, the origin of these two viral isolates needs to more investigation. Most Indian samples are found to classify between G and I clades. One set of thirteen Indian samples (n=13) that belong to the cluster B, which is enriched with European samples as per figure 2, segregate into G clade as they carry D614G mutation in S gene. Another set of thirteen Indian samples (n=13) that belong to the cluster A, which are enriched with Oceania samples as per figure 2, segregate into I clade as they carry V378I mutation in ORF1ab gene. We have also extended our viral clade analysis to recently submitted Indian samples (n=9) from the state of Karnataka. Due to the fact that these samples are low coverage samples, they could not be included in phylogenetic analysis. However, clade analysis revealed three samples from this Karnataka set belongs to G clade, while the status of other samples could not be clearly established (**Supplementary figure 3**). We conclude from this analysis that Indian samples are divided between I and G clades. Thus, these results provide additional confirmation to our earlier findings (as per figure 2) that while one set of Indian samples in cluster A is more similar to Oceania samples, the other set of Indian samples in cluster B is more similar to European samples.

**Figure 3.**
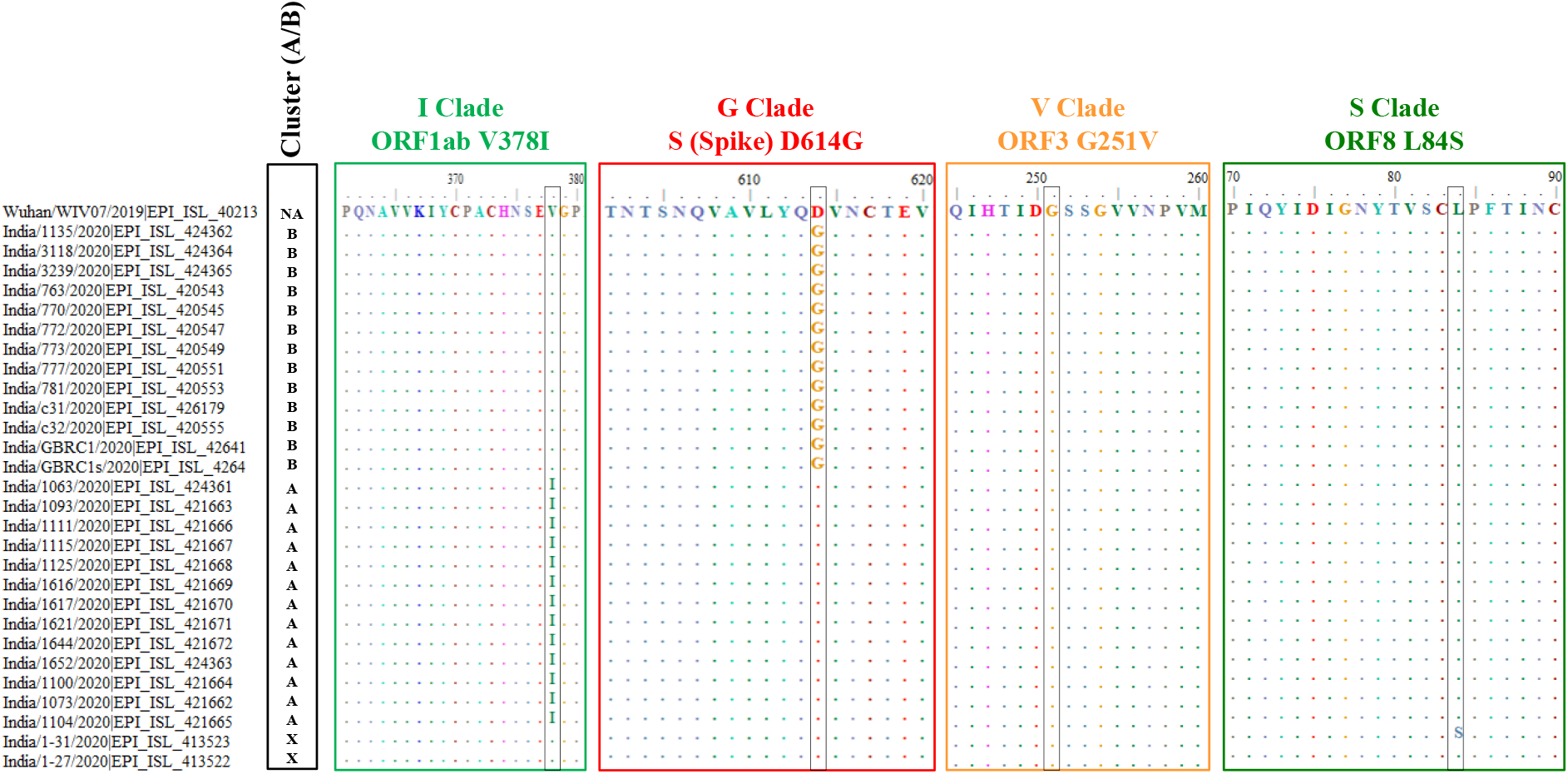
Characterization of clade defining genetic markers of 28 viral sequences from the Indian isolates included in this study. Cluster (A/B) shows the Indian samples belonging to the clusters identified by phylogeny analysis. I clade contains V378I marker in the ORF1ab region, G clade contains D614G marker in the spike protein (S), V clade contains G251V in ORF3 and S clade contains L84S marker in ORF8 region. NA refers to Not applicable; X refers that the samples does not belong either A or B.

Our finding that Indian SARS-CoV-2 isolates belong to different clades may have important consequences with respect to virus transmission rate and virulence, extend of the disease severity and various other aspects of disease pathogenesis. It has been reported that viruses belonging to different clades may differ in their virulence (Brusky, 2020). For example, the G clade viruses carry glycine (G) corresponding to the codon 614 of S protein instead of aspartic acid (D) in other clades. Phylogenetic analysis identified that D614G mutation is originated from ancestral D residue seen in the reference Wuhan virus (Wu et al., 2020). This residue is located very close to glycosylation region of the viral spike protein encoded by S gene (Anderson et al., 2020). It has been proposed that mutations in and around glycosylation region may alter viral spike protein structure and hence the membrane fusion process resulting in varied pathogenicity and transmissibility. Further, the difference in the death rate of COVID-19 patients of East Coast Vs. West Coast of USA is implicated to their difference in their G clade status (Brusky, 2020). There are also many reports that mutations in spike protein of other Coronaviruses alter the virulence (Krueger et al., 2001; Geoghegan and Holmes, 2018; Ontiveros et al., 2003).

The presence of multiple clades of SARS-CoV-2 strains in a population may also have serious implications in the accuracy of diagnostic tests that are being employed worldwide. The diagnostic tests based on detection of antibody or quantifying the viral RNA genome does not appear to distinguish these variations. Consequently, it is important to develop diagnostic kits based on the type(s) of clades prevalent in an area. Indeed, it is reported that the serology based rapid tests are ineffective in detecting COIVD-19 positive cases in India and elsewhere in the world. The variation created by the presence of different viral clades in a population also needs to be considered seriously in developing vaccines. It has been also proposed that relaxation models of social distancing should consider the presence of one or more types of viral clades (Brusky, 2020).

## Conclusions

Collectively, we conclude that the Indian SARS-CoV-2 viruses belong to two major clades (I and G) based on this limited analysis. The probable source of Indian SARS-CoV-2 viruses is countries from Europe and Oceania regions besides Middle East and South Asian regions. The possible spread of the SARS-CoV-2 virus to India through Middle East countries from Europe and Oceania regions cannot be ruled out. Both A and B clusters contain samples from Middle East countries. In addition, these samples appear to split between I and G clades (**Supplementary figure 3**). In the absence of the information related to travel/contact history of Indian patients, more inference and definite conclusions on the possible origin and source could not be made at present. Thus our result also indicate that there is a close connection between source of virus and the countries that are most travelled by Indians. The study also highlights the power of rapid viral genome sequencing and public data sharing to improve the detection and management of pandemic diseases such as COVID-19. It is important to point out that most countries in America, Europe, Oceania and East Asia were quick in supporting the advanced scientific studies on the virus and disease process itself to develop treatment modalities and preventive vaccines. Needless to say that major countries from emerging economies such as Brazil and India should support experimental research on SARS-CoV-2 pathogenesis.

Certainly, our analysis has clear limitations, the most important one being that we were able to analyse only a small number of Indian SARS-CoV-2 genomes while the number of COVID-19 cases increased beyond 20,000. Further, lack of travel history of the patients is another lacuna which makes the conclusions not conclusive. Hence, it is required that more number of Indian isolates of SARS-CoV-2 needs to be sequenced to confirm these results. Nevertheless, our study highlights the need for large-scale community surveillance for SARS-CoV-2 introductions and the spread. More importantly, this work underscores the power of pathogen genomics to identify epidemiological understanding of the virus and the disease.

## Samples and Methods

### Sample Collection

We have collected 28 genome sequences from Indian clinical samples deposited at GISAID. In addition, we also collected another 421 representative genome sequences of samples from USA (75), Europe (80), China (75), East Asia (64), South Asia (41), Oceania (75) and Middle East (11). The viral sequences having complete and high coverage were alone selected. Among Indian viral isolates, seven viral genome sequences that belong to passaged virus through cell lines (Vero CCL81 isolate P1) were excluded in this study. A total of 28 Indian viral genomes were used in this study. However, an additional 9 Indian SARS-CoV-2 genomes with low coverage was used only for clade analysis. Genome accession and sample data information can be found in “**SupplymentaryData**.**xlsx**”.

### Phylogenetic tree analysis

A total of 449 complete genomes were taken for alignment using MAFFT version 7.402 at CIPRES Science Gateway (Miller et al., 2010). Phylogenetic analysis by maximum likelihood (ML) methods was carried out using IQ tree version 1.6.12 (Jana et al., 2016). TIM+F+R2 having lowest BIC score (103964.668) was selected as best substitution model out of 279 substitution model fitted. Analysis was carried out with 10^3 ultrafast bootstrap replicates. The tree file obtained was visualized using Figtree version 1.4.4 (http://tree.bio.ed.ac.uk/software/figtree/).

### Viral clade analysis

A reference Wuhan isolate, Indian and Middle East (Kuwait and Saudi Arabia) whole genome sequences were obtained. Individual genes namely ORF1ab, S, ORF3 and ORF8 were extracted from the whole genome. Obtained genes were aligned using CLUSTAL Omega algorithm (Madeira et al., 2019) and translated to amino acid sequences. The aligned protein coding genes was visualized in BioEdit version 7.2.5 (Hall et al., 1999).

## Data Availability

Data is taken from www.gisaid.org

## Author Approvals

all authors have seen and approved the manuscript, and that it hasn’t been accepted or published elsewhere.

## Acknowledgments

The results published here are in whole or part based upon data generated by Global Initiative on Sharing All Influenza Data (GISAID) (https://www.gisaid.org/). We gratefully acknowledge the authors, originating and submitting laboratories of the sequences from GISAID’s EpiCoV™ Database. KS acknowledges DBT, DST-SERB and ICMR, Government of India for Research grant. Infrastructure support by funding from DST-FIST, IISc-DBT partnership and UGC (Centre for Advanced Studies in Molecular Microbiology) to MCB is acknowledged. KS is a J. C. Bose Fellow of the Department of Science and Technology.

## Abbreviations

SARS-CoV-2: Severe Acute Respiratory Syndrome Coronavirus 2
COVID-19: Coronavirus disease 19
WHO: World Health Organization
GISRS: Global Influenza Surveillance and Response System
GISAID: Global Initiative on Sharing All Influenza Data

## Figure legends

**Supplementary figure 1.**
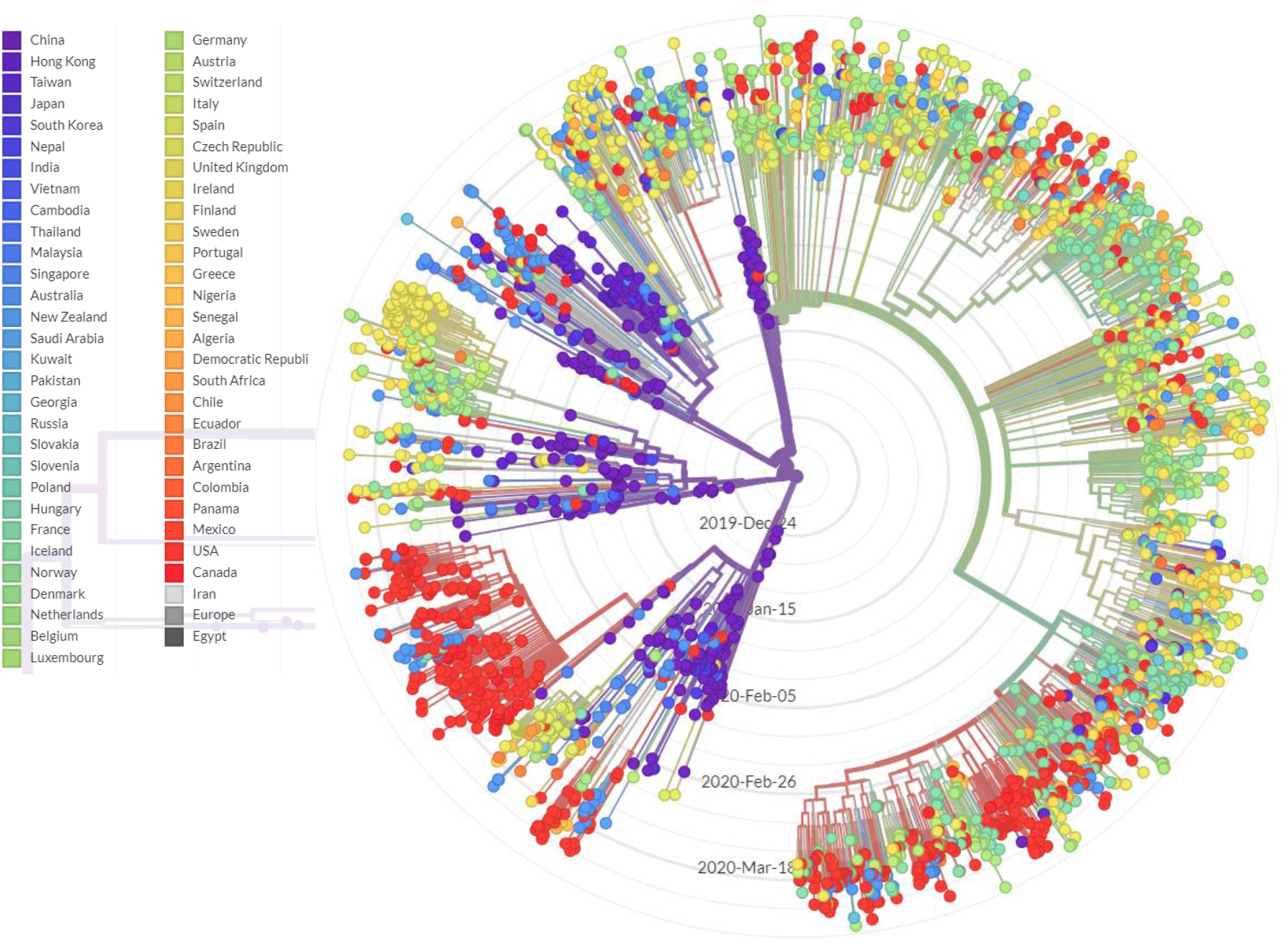
A modified view of Phylogenetic analysis (Radial view) of genome sequences of SARAS-CoV-2 (n=3123) taken from https://www.gisaid.org/epiflu-applications/next-hcov-19-app/. The list of countries from where samples were used is given with their colour code.

**Supplementary figure 2.**
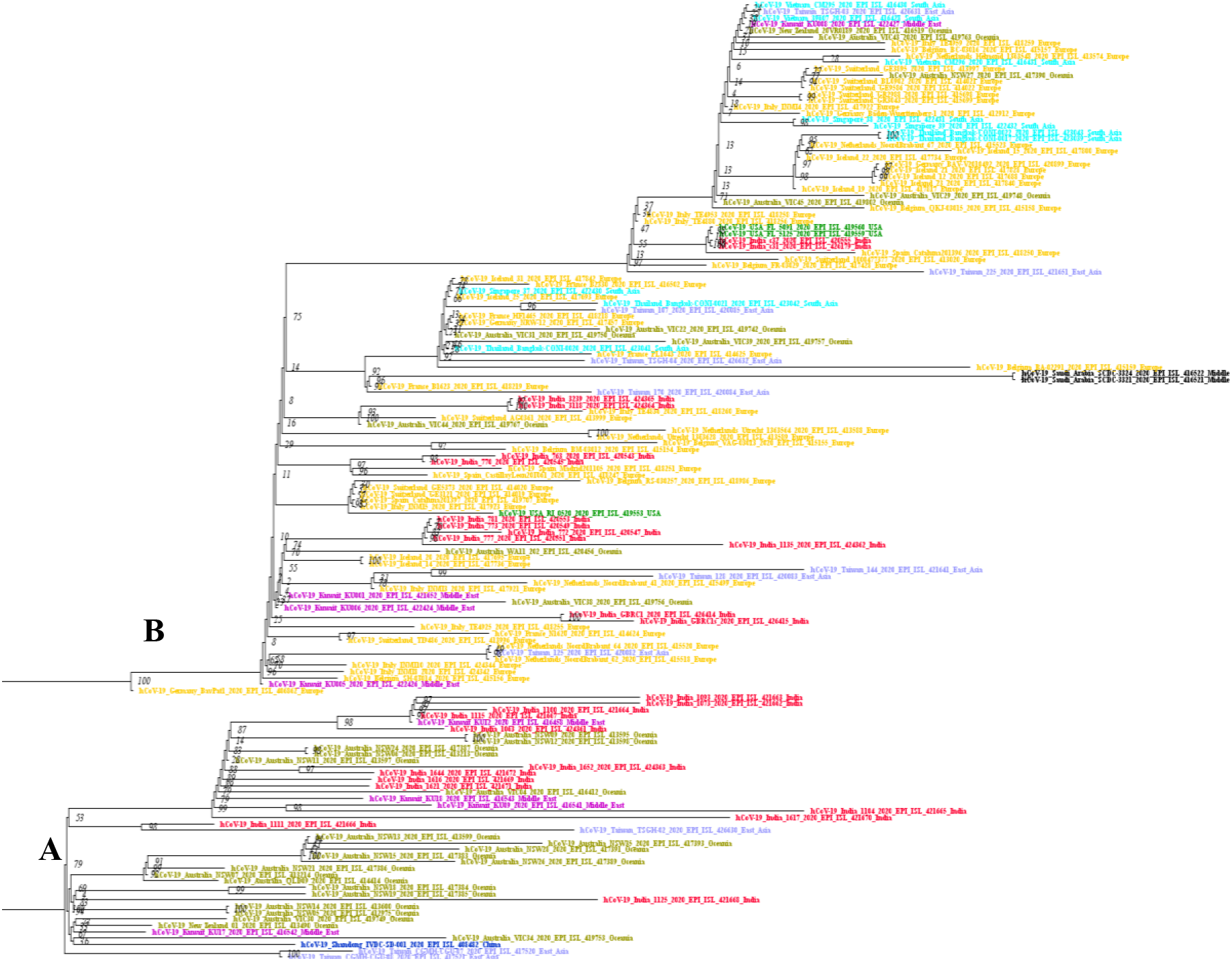
A magnified view of Clusters A and B as per Figure 2 is shown.

**Supplementary figure 3.**
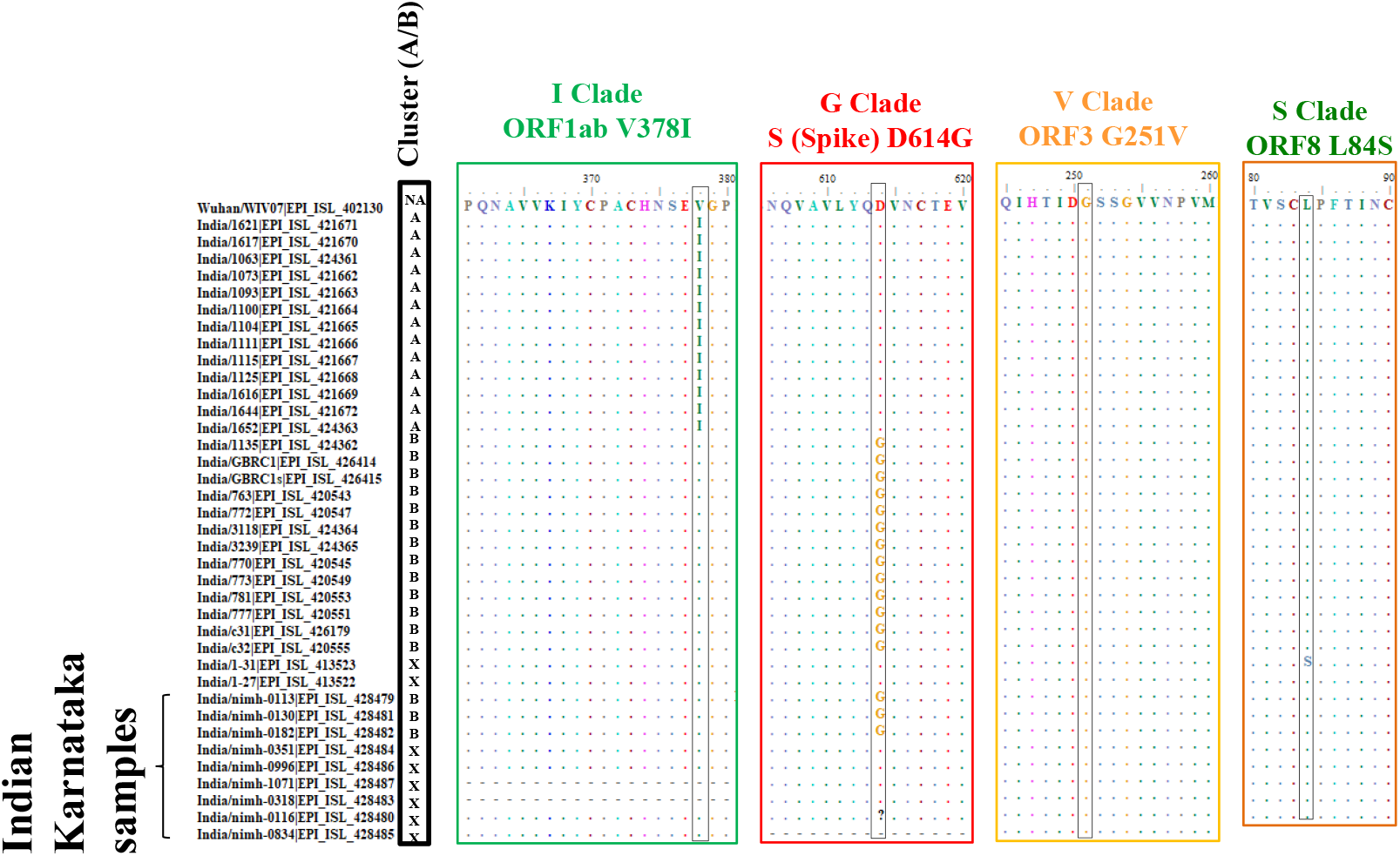
Characterization of clade defining genetic markers of 9 viral sequences from Karnataka, India. Cluster (A/B) shows the Indian samples belonging to the clusters identified by phylogeny analysis. I clade contains V378I marker in the ORF1ab region, G clade contains D614G marker in the spike protein (S), V clade contains G251V in ORF3 and S clade contains L84S marker in ORF8 region. NA refers to Not applicable; X refers that the samples does not belong either A or B.

**Supplementary figure 4.**
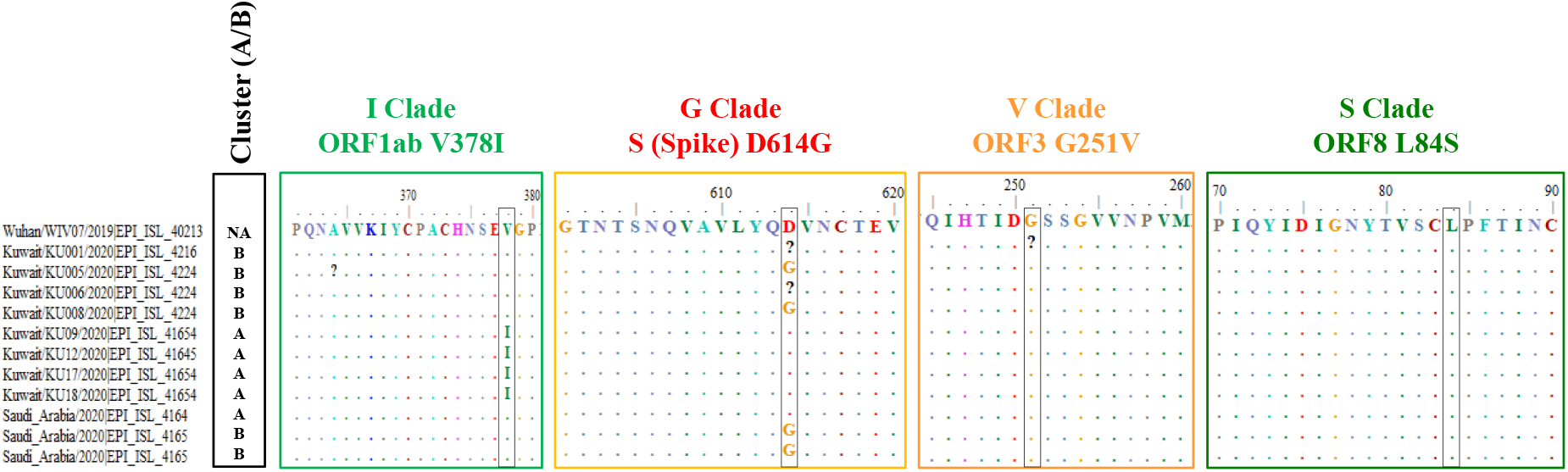
Characterization of clade defining genetic markers of 11 viral sequences from the Middle East (Kuwait/Saudi Arabia) isolates included in this study. Cluster (A/B) shows the Middle East samples belonging to the clusters identified by phylogeny analysis. I clade contains V378I marker in the ORF1ab region, G clade contains D614G marker in the spike protein (S), V clade contains G251V in ORF3 and S clade contains L84S marker in ORF8 region. NA refers to Not applicable.

